# The Impact of Probiotics on Wellbeing: An Open-Label study during the Winter in Healthcare Workers

**DOI:** 10.1101/2025.08.19.25333967

**Authors:** Clare Wright, Catherine Goodwin, Daniel John, Daryn Michael, Niall Coates, Tom Webberley, Sue Plummer, Keri Turner

**Affiliations:** Cwm Taf Morgannwg University Health Board, Pontypridd, United Kingdom; Wales Ambulance services NHS Trust, Wales, United Kingdom; Cultech Limited, Unit 2 Christchurch Road, Baglan Industrial Park, Port Talbot, United Kingdom; Seastorm Ltd, Cardiff, United Kingdom

**Keywords:** probiotic, healthcare worker, wellbeing, real world evidence

## Abstract

Employment within healthcare settings can place a significant strain on the general wellbeing of staff, particularly during the winter. There is a link between health and wellbeing and the composition of the gut microbiota and daily supplementation with probiotics has been shown to stabilise/modulate the gut microbiota which may help support the health and wellbeing of healthcare workers.

In this exploratory, single-arm, open-label, remotely-conducted study, National Health Service employees in Wales received a daily dose of probiotic for 8 weeks over the winter season and those volunteering to take part reported their perception of quality of life including sleep quality, energy levels and mood and their physical discomforts including bloating and muscle ache at weekly intervals throughout the intervention period (ClinicalTrials.gov: NCT05968209).

Compared to the start of the study, their sleep quality significantly improved by 34.2%, their energy levels by 29.2% and their overall mood by 24.3% after 8 weeks of probiotic supplementation. Their general wellbeing had significantly improved by 16%. The prevalence of bloating decreased significantly from 75% at the start of the study to 42% by the study end, and muscle aches fell from 76% to 45%.

The findings indicate that the wellbeing of healthcare workers over the winter months improved whilst receiving daily probiotic supplementation. Further work is required in a placebo-controlled, randomised, double-blind study.

## 1. Introduction

Employee wellbeing is important both for people and organisations and promoting wellbeing can help to prevent stress and create positive working environments, improve employee engagement and optimise overall staff performance^1^. Working within healthcare settings can place a significant strain on employee wellbeing due to ever increasing numbers of patients fuelling heavy workloads and stress, whilst irregular shift patterns can disrupt circadian rhythms^2^ and frontline staff also face a heightened risk of communicable infections^3^. The rates of absenteeism are often high within the healthcare workforce especially during the winter when there are the added pressures of seasonal illnesses (colds and flu) and seasonal affective disorder^4^.

One approach optimise employee wellbeing is to consider options that overall encourage workplace health promotion and there is growing awareness of the link between health and the composition and functionality of the human gut microbiota when in a stable, balanced state^5^. The gut microbiota - a collective term for the trillions of microorganisms residing in the gastrointestinal tract – can play important roles in many physiological processes including metabolism, immunity and cognition^5, 6^. Disturbances to the composition of the gut microbiota can occur as the result of poor diet, lifestyle and the environment and are strongly associated with deficits in host health and wellbeing^7^. Providing opportunities to optimise the structure and function of the gut microbiota to prevent its disruption may be one possible approach to support the health and wellbeing of the host^5^.

Probiotics are defined as ‘live microorganisms that, when administered in adequate amounts, confer a health benefit on the host’^8^ and they have been shown to provide a means of stabilising and optimising the gut microbiota^9^. A number of studies have assessed the impact of supplementation with probiotics on the wellbeing of healthcare workers, particularly those who are patient-facing^10–13^, reporting better sleep quality and reduced incidence of constipation^13^ along with the prevention of respiratory tract infections^12^.

The Lab4P probiotic has been shown to improve wellbeing and mental health and reduce incidence rates of stomach pains, bloating, muscle aches and headaches^14–17^. In an earlier workforce based voluntary and anonymised Lab4P pilot study, there were improvements in several wellbeing measures including sleep quality, energy levels and mood and also reductions in the incidence rates of common gastrointestinal and respiratory symptoms^18^.

When running human intervention studies there is a need to minimise the disruptive impacts on the participants who might be required to adhere to strict study conditions and be burdened with repeated visits to a study centre for monitoring and the completion of paper based questionnaires. Real-World Evidence (RWE) human intervention studies are gaining popularity as they offer a means of assessment during routine, everyday life, with minimal impact from the study conditions^19^. Advancements in digital technologies are aiding RWE studies allowing for fully remote recruitment and data capture thus lowering the demands placed on study participants^20^.

In this study, we utilised a smartphone based app (*Trialflare*) to conduct an exploratory, single-arm, 8-week, open-label study in healthcare workers during the winter. The objectives were to gauge willingness of people to engage in the study and their levels of retention and compliance and to evaluate the impact of daily Lab4P supplementation on participant wellbeing.

## 2. Methods

### 2.1. Design, Approval, Eligibility & Recruitment

An exploratory, single-arm, open-label, 8-week probiotic intervention study conducted in accordance with the principles of the Declaration of Helsinki with ethical approval from the Health Research Authority and Health and Care Research Wales (reference 23/HCRW/0033). The study was prospectively registered with ClinicalTrials.gov (NCT05968209).

Eligible participants were aged 18 to 60, employed by NHS Wales at the Cwm Taf Morgannwg University Health Board (CTM UHB) or the Welsh Ambulance Services University NHS Trust (WAST), fluent in English, willing/able use their personal smartphone as part of the study, and willing to maintain their usual lifestyle for the duration of the study. Exclusion criteria included any known immunodeficiency, allergy to zinc, and current or recent probiotic use.

The study was advertised via posters, email and social media and directed anyone interested to the study website. The website invited those interested to review the study information sheet and eligibility criteria before deciding whether to provide informed consent on-line (eConsent). Consenting participants were advised (via text message) where they could collect their probiotic product. They were also encouraged (but not obliged) to download the *Trialflare* App (Seastorm Ltd, Cardiff, UK) which is an easy-to-use platform for collecting data securely and anonymously from the participants at regular intervals throughout the study.

### 2.2. Study Intervention

The probiotic product (Lab4P) comprised capsules containing *Lactobacillus acidophilus* strains CUL-60 (NCIMB (National Collection of Industrial, Food and Marine Bacteria) 30157), and CUL-21 (NCIMB 30156), *Lactiplantibacillus plantarum* CUL-66 (NCIMB 30280), *Bifidobacterium bifidum* CUL-20 (NCIMB 30153), and *Bifidobacterium animalis* subsp*. lactis* CUL-34 (NCIMB 30172) totalling 50 billion colony-forming unit (CFU), Vitamin D (10 µg), Vitamin C (80 mg), and Zinc (10 mg). Capsules were packed in induction-sealed polyethylene pots (60 capsules per pot). The participants took one capsule daily (with or following food but not with hot drinks) and were advised to store the pots in a refrigerator.

### 2.3. Study Outcomes & Data collection

The outcomes were compliance to the study (interest, data submission and product intake), changes in quality of life, gastrointestinal and other common discomforts and absenteeism. The participant questionnaires were completed remotely on the *Trialflare* app. Participants received weekly alerts via the *Trialflare* app, reminding them to take the study product and complete the questionnaires.

### 2.4. Demographics

Completed at baseline and recording participant age (years), sex assigned at birth (*male/female/prefer not to say*), approximate weight (kilograms or stones/lbs), approximate height (centimetres or feet/inches) and place of work (*CTM UHB* or *WAST*).

### 2.5. Participant questionnaire

Completed at baseline and at weekly intervals throughout the intervention period, a recall questionnaire of the previous 7 days recording:

*i)* the participants *Quality of life* across 5 domains: General Wellbeing, State of Mood, State of Health, State of Energy and Sleep Quality each rated on a 10 point Likert scale between 1 = ‘Worst’ and 10 = ‘Best’^16^.
*ii)* the number of days experiencing *Physical discomfort* including gastrointestinal related discomforts (constipation, diarrhea, bloating, stomach pain and/or indigestion) and other common discomforts (coughing, sore throat, headache, muscle ache and/or chest wheezing).
*iii)* the number of days that the participant was required to work and absent from work (with reason: *illness/prefer not to say/other/not applicable*).
*iv)* whether the participant was still taking the study product (*yes/no*).

### 2.6. Statistical Analysis

Statistical analysis was performed on an intention-to-treat basis for all participants providing data during the study. Quality of life scores were analysed using Mixed-Effects Models (MEMs) with participant as the random effect, and baseline, time-point and product compliance as the fixed effects. Results are reported as adjusted least square mean (LSM) change from baseline. Occurrence of physical discomforts were analysed using a Binomial Mixed-Effects Model (BMEM) with participant as the random effect and baseline, time-point and study product compliance as the fixed effects. Results are presented as the proportion of participants reporting symptom(s) each week. Absence rates due to illness (%) were calculated as *((days absent with illness)/(days required to work))*100* and analysed using Kruskal-Wallis test with Dunn’s *post-hoc* analysis (GraphPad Prism, Version 10.4.1). Values of *p* were considered statistically significant when ≤0.05.

## 3. Results

The study flow diagram is shown in Figure 1. A total of 390 individuals signed up and provided eConsent between 02/09/2024 and 19/12/2024 and of these, 174 participants collected the study product and registered with the *Trialflare* app. Ninety-seven participants recorded data throughout the 8-week intervention period. Participant-reported product compliance exceeded 90%. No serious adverse events were reported during the study.

**Figure 1:**
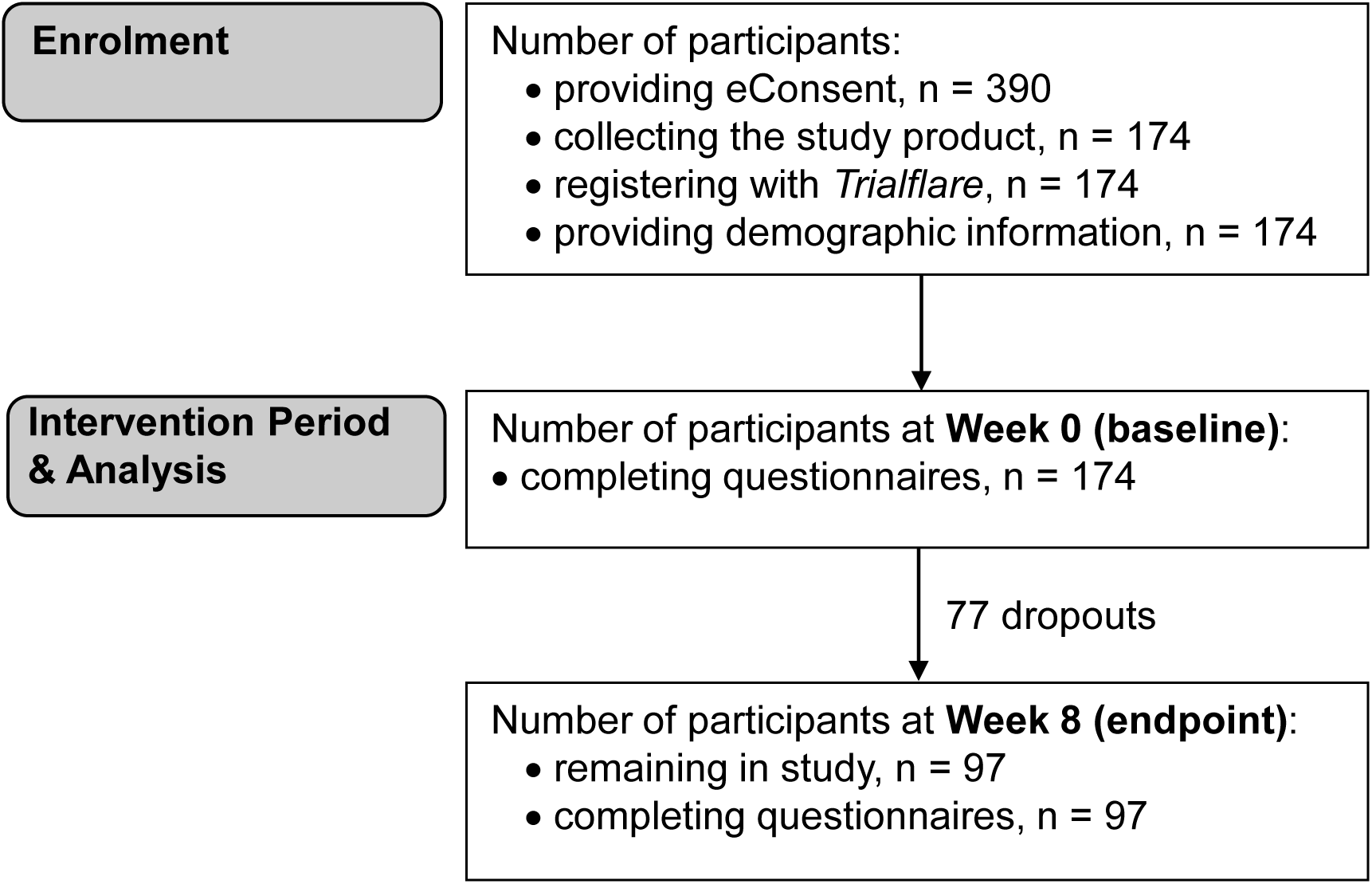
Study flow diagram. Number of participants at Week 1=159, Week 2=149, Week 3=139, Week 4=126, Week 5=111, Week 6=102, Week 7=98 and Week 8=97.

The baseline characteristics of the study population are shown in Table 1. The majority of participants were female (83%) and/or employed by CTM UHB (67%). Based on participant assessments of their BMI, 42% of the total population were obese.

**Table 1.**
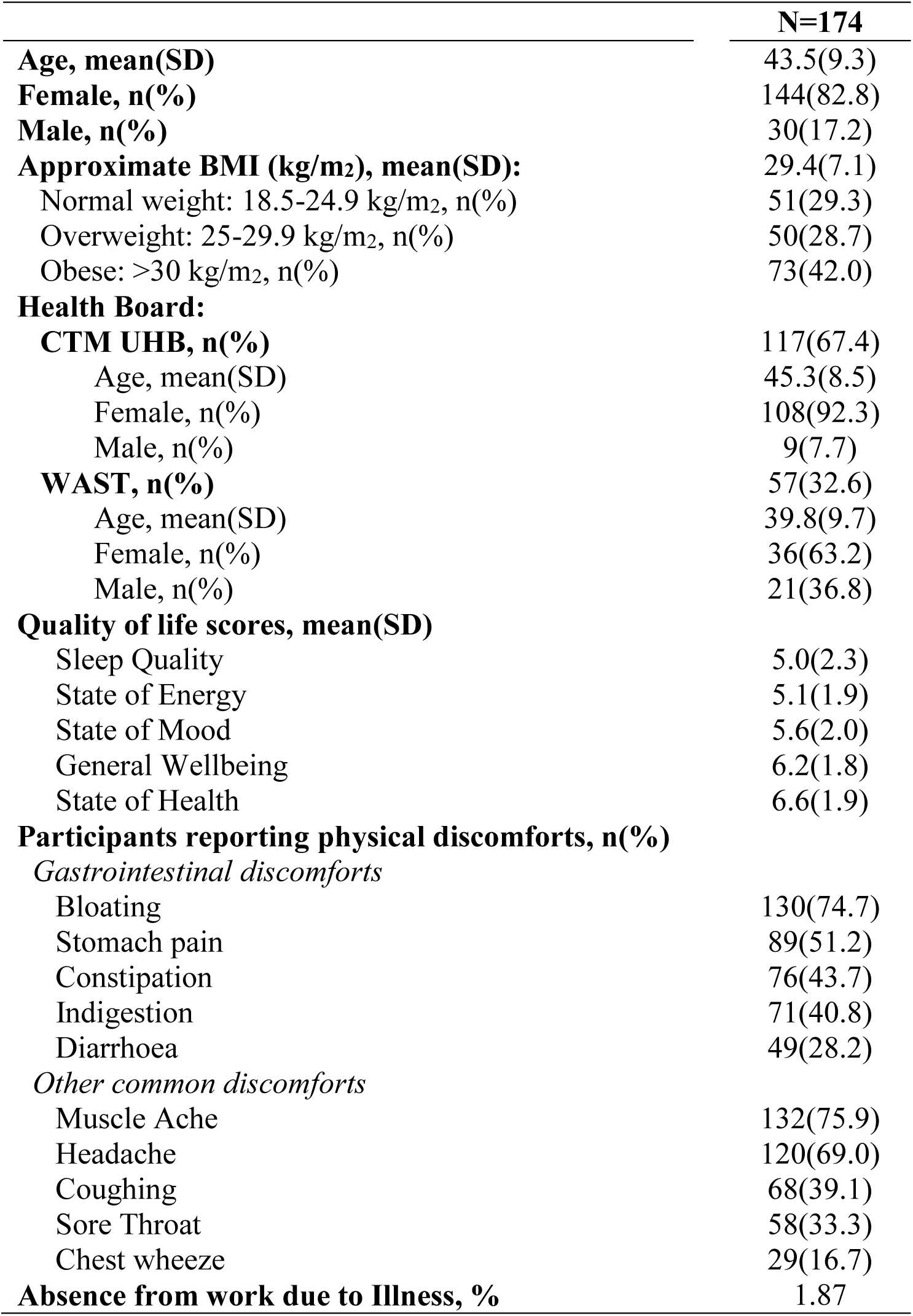
Baseline characteristics.

|  | **N=174** |
| --- | --- |
| **Age, mean(SD)** | 43.5(9.3) |
| **Female, n(%)** | 144(82.8) |
| **Male, n(%)** | 30(17.2) |
| **Approximate BMI (kg/m<sup>2</sup>), mean(SD):** | 29.4(7.1) |
| Normal weight: 18.5-24.9 kg/m<sup>2</sup>, n(%) | 51(29.3) |
| Overweight: 25-29.9 kg/m<sup>2</sup>, n(%) | 50(28.7) |
| Obese: >30 kg/m<sup>2</sup>, n(%) | 73(42.0) |
| **Health Board:** |  |
| **CTM UHB, n(%)** | 117(67.4) |
| Age, mean(SD) | 45.3(8.5) |
| Female, n(%) | 108(92.3) |
| Male, n(%) | 9(7.7) |
| **WAST, n(%)** | 57(32.6) |
| Age, mean(SD) | 39.8(9.7) |
| Female, n(%) | 36(63.2) |
| Male, n(%) | 21(36.8) |
| **Quality of life scores, mean(SD)** |  |
| Sleep Quality | 5.0(2.3) |
| State of Energy | 5.1(1.9) |
| State of Mood | 5.6(2.0) |
| General Wellbeing | 6.2(1.8) |
| State of Health | 6.6(1.9) |
| **Participants reporting physical discomforts, n(%)** |  |
| *Gastrointestinal discomforts* |  |
| Bloating | 130(74.7) |
| Stomach pain | 89(51.2) |
| Constipation | 76(43.7) |
| Indigestion | 71(40.8) |
| Diarrhoea | 49(28.2) |
| *Other common discomforts* |  |
| Muscle Ache | 132(75.9) |
| Headache | 120(69.0) |
| Coughing | 68(39.1) |
| Sore Throat | 58(33.3) |
| Chest wheeze | 29(16.7) |
| **Absence from work due to Illness, %** | 1.87 |

### 3.1. Quality of life

At baseline, the worst (lowest) scoring quality of life domain was Sleep Quality, followed by State of Energy, State of Mood, General Wellbeing and State of Health (Table 1). Changes in scores from baseline during the 8-week probiotic intervention period are shown in Figure 2 (details in Supplementary Table S1). There were significant improvements in Sleep Quality, State of Energy and State of Mood throughout, culminating in a 34.2% improvement in Sleep Quality (*p*<0.0001; Figure 2A), a 29.2% improvement in State of Energy (*p*<0.0001; Figure 2B) and a 24.3% improvement in State of Mood (*p*<0.0001; Figure 2C) at Week 8. For General Wellbeing, significant improvements were observed from Week 3 (7.4%, *p*=0.0073, Figures 2D) to Week 8 (16%, *p*<0.0001) and for State of Health, scores had significantly improved from Week 4 (7.1%, *p*=0.0078, Figures 2E) through to Week 8 (10.1%, *p*=0.0004).

**Figure 2.**
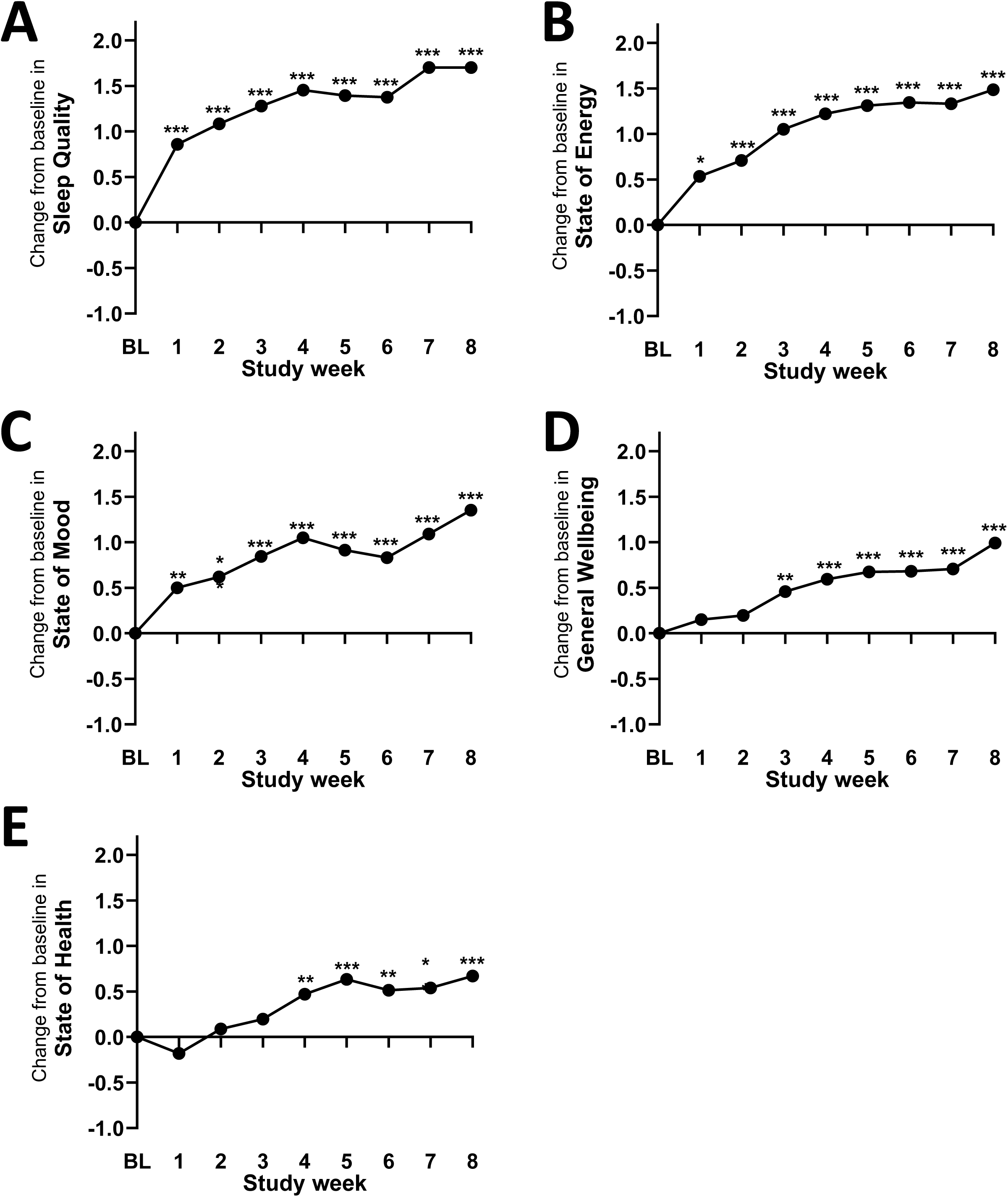
Changes from baseline score in (A) Sleep Quality, (B) State of Energy, (C) State of Mood, (D) General Wellbeing and (E) State of Health during the study. Data presented are least square mean change from baseline for n=159 at Week 1, n=147 at Week 2, n=137 at Week 3, n=125 at Week 4, n=110 at Week 5, n=102 at Week 6, n= 98 at Week 7 and n=97 at Week 8. Values of p were determined by mixed-effect modelling where *p≤0.05, **p≤0.01, and ***p≤0.001 vs. baseline (BL).

### 3.2. Physical Discomfort

Figure 3 shows the proportion of participants reporting physical discomfort during the study (details in Supplementary Table S2). At baseline, 74.7% of participants reported bloating (Figure 3A), which significantly reduced to 41.6% at Week 4 (*p*=0.0193) and 36.1% at Week 8 (*p*=0.0013). Significant reductions from baseline were also observed for the proportion of participants with stomach pain, initially reported by 51.1% of participants (Figure 3B) but declining to approximately half of that from Week 4 (24%, *p*=0.0167), Week 7 (24.5%, *p*=0.0473) and Week 8 (25.8%, *p*=0.0714). Constipation was reported by 43.7% of participants at baseline (Figure 3C) and decreased significantly to 20.9% by Week 5 (*p*=0.0259) and was 21.7% at Week 8 (*p*=0.0584). Indigestion and diarrhoea incidences did not change (Supplementary Figure S1).

**Figure 3.**
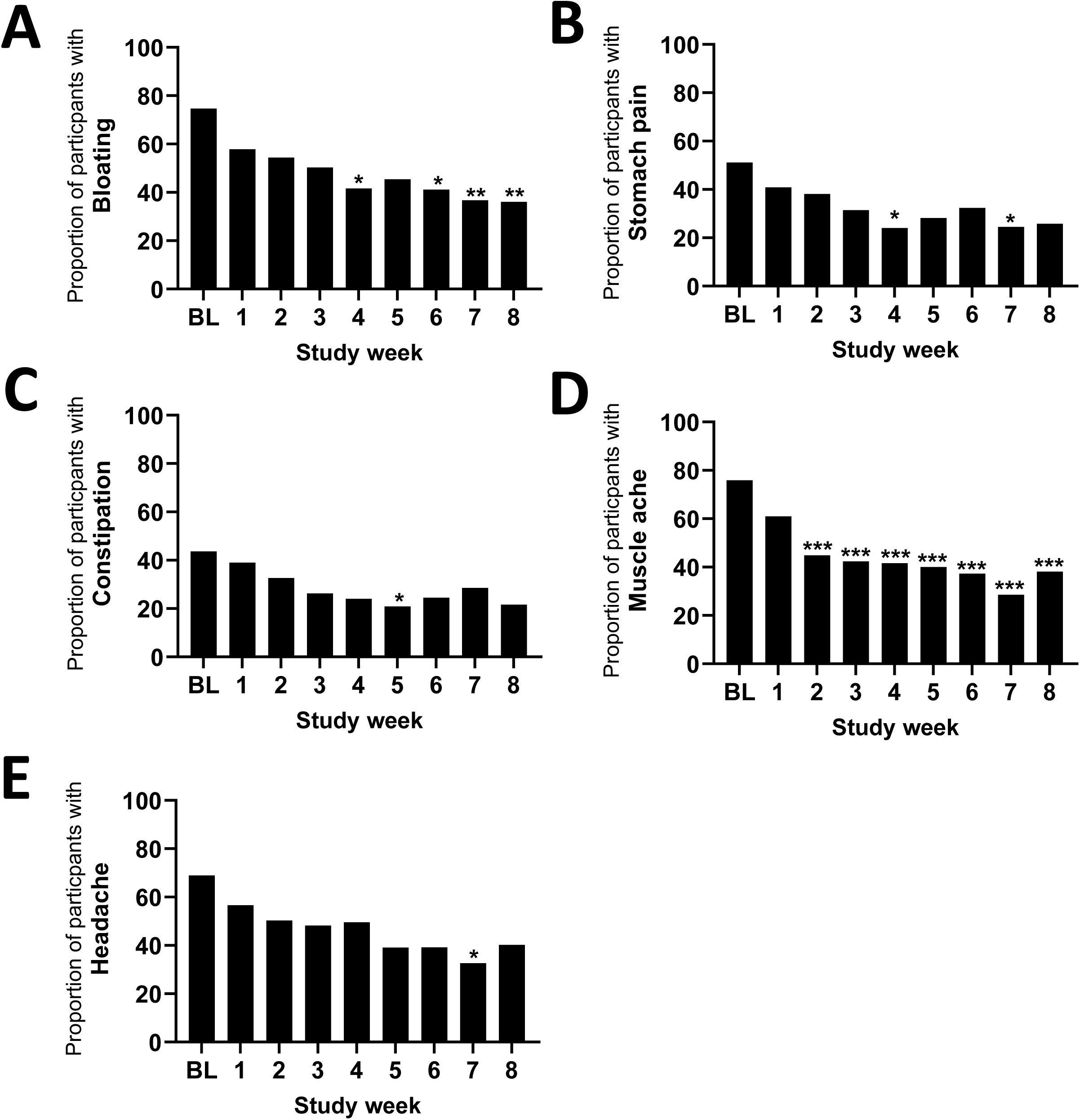
Incidence of participants reporting (A) Bloating, (B) Stomach pain, (C) Constipation, (D) Muscle Ache and (E) Headache over the duration of the study. Data are presented as proportion (%) of participants reporting symptoms for n=159 at Week 1, n=147 at Week 2, n=137 at Week 3, n=125 at Week 4, n=110 at Week 5, n=102 at Week 6, n= 98 at Week 7 and n=97 at Week 8. Values of *p* were determined by binomial mixed-effect modelling where \**p*≤0.05, \*\**p*≤0.01, and \*\*\**p*≤0.001 vs. baseline (BL).

For the non-GI-related discomfort, muscle ache was the most commonly reported by 75.9% of the participants at baseline, decreasing to 44.9% (*p*=0.0001, Figure 3D) after 2 weeks and to 38.1% by endpoint (*p*<0.0001). The proportion of participants reporting headache also significantly reduced during the study, from 69.0% at baseline to 32.7% at Week 7 (*p*=0.0156, Figure 3E). No significant changes were observed for coughing, sore throat or chest wheeze (Supplementary Figure S1).

### 3.3. Absence Rates

Weekly absence rates due to illness are shown in Figure 4. Changes in absteneeism during the intervention period were not statistically significant.

**Figure 4.**
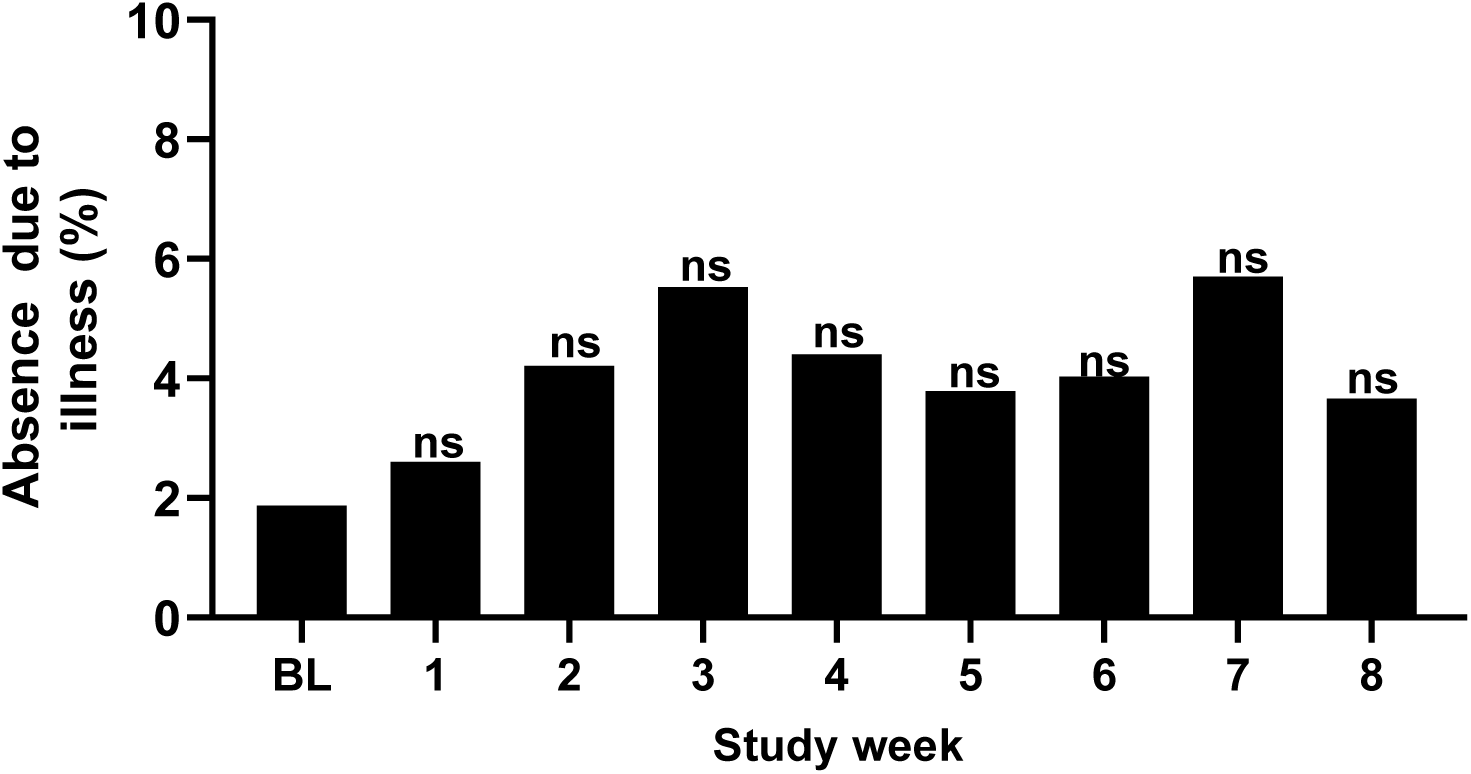
Absence due to illness. Data are presented as mean % absence for n=171 at baseline (BL) of participants reporting symptoms for n=150 at Week 1, n=134 at Week 2, n=130 at Week 3, n=118 at Week 4, n=104 at Week 5, n=198 at Week 6, n= 92 at Week 7 and n=91 at Week 8. Values of *p* were determined using Kruskal-Wallis test with Dunn’s post hoc where ‘ns’ indicates not significant vs. baseline.

## 4. Discussion

One hundred and seventy-four healthcare workers enrolled in the study using the remote eConsent and data submission process for this exploratory, single-arm, open-label study. During the 8-week intervention period there were significant improvements in the following quality of life measures: Sleep Quality, State of Energy and State of Mood, alongside reduced incidence of physical discomforts including bloating, stomach pain, constipation, muscle ache and headaches in this predominantly female cohort.

From the baseline scores for the Sleep Quality and State of Energy questions, it appeared that sleep-related issues and fatigue were prevalent within the study cohort, and these elements showed the greatest improvement over the duration of the study. The results from a meta-analysis of participants with sleep problems found that probiotic supplementation can improve sleep quality^21^ and there is also evidence supporting the fatigue reducing capabilities of probiotics^22^. In previous placebo controlled studies with the Lab4P probiotic there have been improvements in both sleep quality and energy levels^16, 17^ and a study with *Lactobacillus delbrueckii* ssp. *bulgaricus* OLL1073R-1, improved sleep and vitality in a cohort of female healthcare worker^13^. Sleep deprivation is often accompanied by muscle aches and headaches^23, 24^ and incidence rates of both symptoms were significantly reduced in the healthcare workers. These outcomes are in line with the results from a previous winter based Lab4P probiotic study in overweight and obese adults^25^.

Probiotics are becoming recognised for their ability to improve the mental health of the host, via modulation of the gut brain axis^26^. The State of Mood of the healthcare workers in our study was improved whilst taking the Lab4P supplement, and similar improvements in mood scores were seen in a study with Lab4P in overweight and obese adults^16^ and in a study in a cohort of women with irritable bowel disease there were reductions in anxiety and depression^14^. These findings provide support for a potential role for the Lab4P probiotic in the functioning of the gut-brain-axis.

General Wellbeing and State of Health were reported by the participants and both measures significantly improved during the period of supplementation. In a study with Lab4P with women of similar age to those in this female dominated study there were significant (17 %) improvements in ‘overall wellbeing’^17^, adding further support to the likely contribution of the Lab4P probiotic to the benefits observated. In the study with female healthcare workers receiving *Lactobacillus delbrueckii* ssp. *bulgaricus* OLL1073R-1, there was an improvement in ‘general health’ which occurred alongside significant reductions in the incidence of constipation^13^.

Gastrointestinal (GI) problems are known to represent a problem for healthcare workers^27^ and particularly for females^28^ which is attributed to a number of factors including the stressful working environment that can disrupt the balance of the gut-brain-axis^29^. In this study, there were high rates of bloating and stomach pain at the start which had significantly reduced after 8 weeks and these improvements occurred alongside reduced incidence of constipation. Healthy adults and people suffering with Irritable Bowel Syndrome who have received the probiotic showed reductions in gastrointestinal discomfort and/or constipation^14,15, 30^.

Health and wellbeing issues in healthcare workers have been linked with systemic inflammation and oxidative stress^31, 32^. Some probiotic bacteria have anti-inflammatory and anti-oxidative capabilities^33, 34^ and the Lab4P product has been shown to reduce circulating levels of pro-inflammatory interleukin(IL-6) levels^15, 35^, induce anti-inflammatory Transforming Growth Factor (TGF)-β and IL-10 in peripheral blood mononuclear cells extracted from healthy volunteers^36, 37^ and protect the viability of neuronal cells exposed to oxidative stress^38^.

Absence from work due to illness was recorded throughout the intervention period. Throughout the period of supplementation, no significant changes in absenteeism were noted despite the trial taking place during the winter period when increased absenteeism would be anticipated^4^. Absence rates were 4-5% whereas published absence rates within CTM UHB and WAST averaged ∼7.5% during the period of our study^39^.

In summary, during the period when the healthcare workers were receiving daily Lab4P probiotic supplementation there were significant improvements quality of life and significant reductions in common discomforts including bloating, stomach pain, constipation, muscle ache and headache. The outcomes from this exploratory study provide strong support for a powered, placebo-controlled, randomised, double-blind, study.

## Data Availability

All data produced in the present study are available upon reasonable request to the authors

