## Supplementary Information for "The Impact of Probiotics on Wellbeing: An Open-Label study during the Winter in Healthcare Workers"

**Supplementary Table S1:** Changes from baseline in Quality of life scores

|  | **Week** | **n** | **Change from baseline^#^** | **Lower 95% CI** | **Upper 95% CI** | **% Change from baseline** | ***p* value** |
| --- | --- | --- | --- | --- | --- | --- | --- |
| **Sleep Quality** | 1 | 159 | 0.86 | 0.45 | 1.27 | 17.26 | <0.0001 |
|  | 2 | 147 | 1.08 | 0.68 | 1.49 | 21.76 | <0.0001 |
|  | 3 | 137 | 1.28 | 0.88 | 1.67 | 25.68 | <0.0001 |
|  | 4 | 125 | 1.46 | 1.05 | 1.86 | 29.22 | <0.0001 |
|  | 5 | 110 | 1.39 | 0.99 | 1.80 | 28.00 | <0.0001 |
|  | 6 | 102 | 1.37 | 0.96 | 1.79 | 27.61 | <0.0001 |
|  | 7 | 98 | 1.70 | 1.28 | 2.12 | 34.18 | <0.0001 |
|  | 8 | 97 | 1.70 | 1.29 | 2.11 | 34.17 | <0.0001 |
| **State of Energy** | 1 | 159 | 0.53 | 0.13 | 0.94 | 10.51 | 0.0103 |
|  | 2 | 147 | 0.71 | 0.30 | 1.12 | 13.97 | 0.0006 |
|  | 3 | 137 | 1.05 | 0.65 | 1.45 | 20.71 | <0.0001 |
|  | 4 | 125 | 1.22 | 0.82 | 1.63 | 24.07 | <0.0001 |
|  | 5 | 110 | 1.31 | 0.91 | 1.72 | 25.84 | <0.0001 |
|  | 6 | 102 | 1.35 | 0.92 | 1.77 | 26.51 | <0.0001 |
|  | 7 | 98 | 1.33 | 0.90 | 1.77 | 26.23 | <0.0001 |
|  | 8 | 97 | 1.49 | 1.07 | 1.90 | 29.24 | <0.0001 |
| **State of Mood** | 1 | 159 | 0.50 | 0.13 | 0.87 | 8.98 | 0.0082 |
|  | 2 | 147 | 0.62 | 0.25 | 0.99 | 11.12 | 0.0011 |
|  | 3 | 137 | 0.84 | 0.47 | 1.22 | 15.17 | <0.0001 |
|  | 4 | 125 | 1.05 | 0.68 | 1.42 | 18.84 | <0.0001 |
|  | 5 | 110 | 0.91 | 0.53 | 1.30 | 16.41 | <0.0001 |
|  | 6 | 102 | 0.83 | 0.43 | 1.23 | 14.93 | <0.0001 |
|  | 7 | 98 | 1.09 | 0.70 | 1.48 | 19.59 | <0.0001 |
|  | 8 | 97 | 1.35 | 0.97 | 1.74 | 24.29 | <0.0001 |
| **General Wellbeing** | 1 | 159 | 0.15 | -0.19 | 0.49 | 2.43 | 0.3891 |
|  | 2 | 147 | 0.20 | -0.14 | 0.54 | 3.16 | 0.2580 |
|  | 3 | 137 | 0.46 | 0.12 | 0.80 | 7.40 | 0.0073 |
|  | 4 | 125 | 0.60 | 0.27 | 0.92 | 9.59 | 0.0004 |
|  | 5 | 110 | 0.68 | 0.34 | 1.01 | 10.88 | 0.0001 |
|  | 6 | 102 | 0.68 | 0.33 | 1.04 | 10.97 | 0.0002 |
|  | 7 | 98 | 0.71 | 0.34 | 1.07 | 11.37 | 0.0001 |
|  | 8 | 97 | 0.99 | 0.64 | 1.35 | 15.99 | <0.0001 |
| **State of Health** | 1 | 159 | -0.18 | -0.54 | 0.18 | -2.72 | 0.3222 |
|  | 2 | 147 | 0.09 | -0.27 | 0.44 | 1.32 | 0.6290 |
|  | 3 | 137 | 0.20 | -0.16 | 0.56 | 2.98 | 0.2826 |
|  | 4 | 125 | 0.47 | 0.12 | 0.82 | 7.11 | 0.0078 |
|  | 5 | 110 | 0.63 | 0.28 | 0.98 | 9.58 | 0.0004 |
|  | 6 | 102 | 0.51 | 0.14 | 0.88 | 7.77 | 0.0065 |
|  | 7 | 98 | 0.54 | 0.18 | 0.90 | 8.15 | 0.0037 |
|  | 8 | 97 | 0.67 | 0.30 | 1.04 | 10.12 | 0.0004 |

^#^Least square means

Abbreviations: CI, confidence interval

**Supplementary Table S2:** The proportions of participants reporting physical discomforts

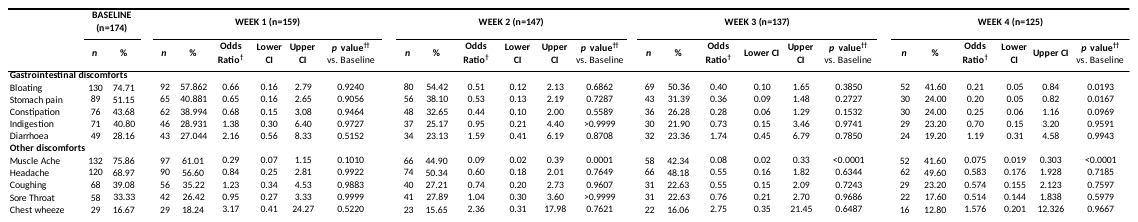

^†^Representing the odds of reporting a symptom compared to baseline: An Odds Ratio(OR) >1 indicates that participants were more likely to report a symptom whereas an OR <1 indicates they were less likely

^††^Binomial mixed-effects models (MEM) with participant as the random effect and compliance to product and baseline score as the fixed effects

**Supplementary Table S2:** Continued….

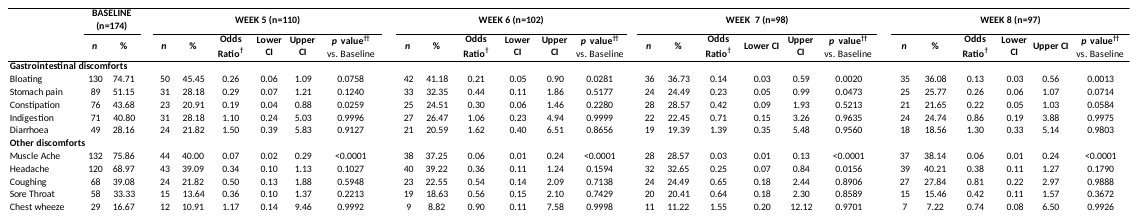

^†^Representing the odds of reporting a symptom compared to baseline: An Odds Ratio(OR) >1 indicates that participants were more likely to report a symptom whereas an OR <1 indicates they were less likely

^††^Binomial mixed-effects models (MEM) with participant as the random effect and compliance to product and baseline score as the fixed effects

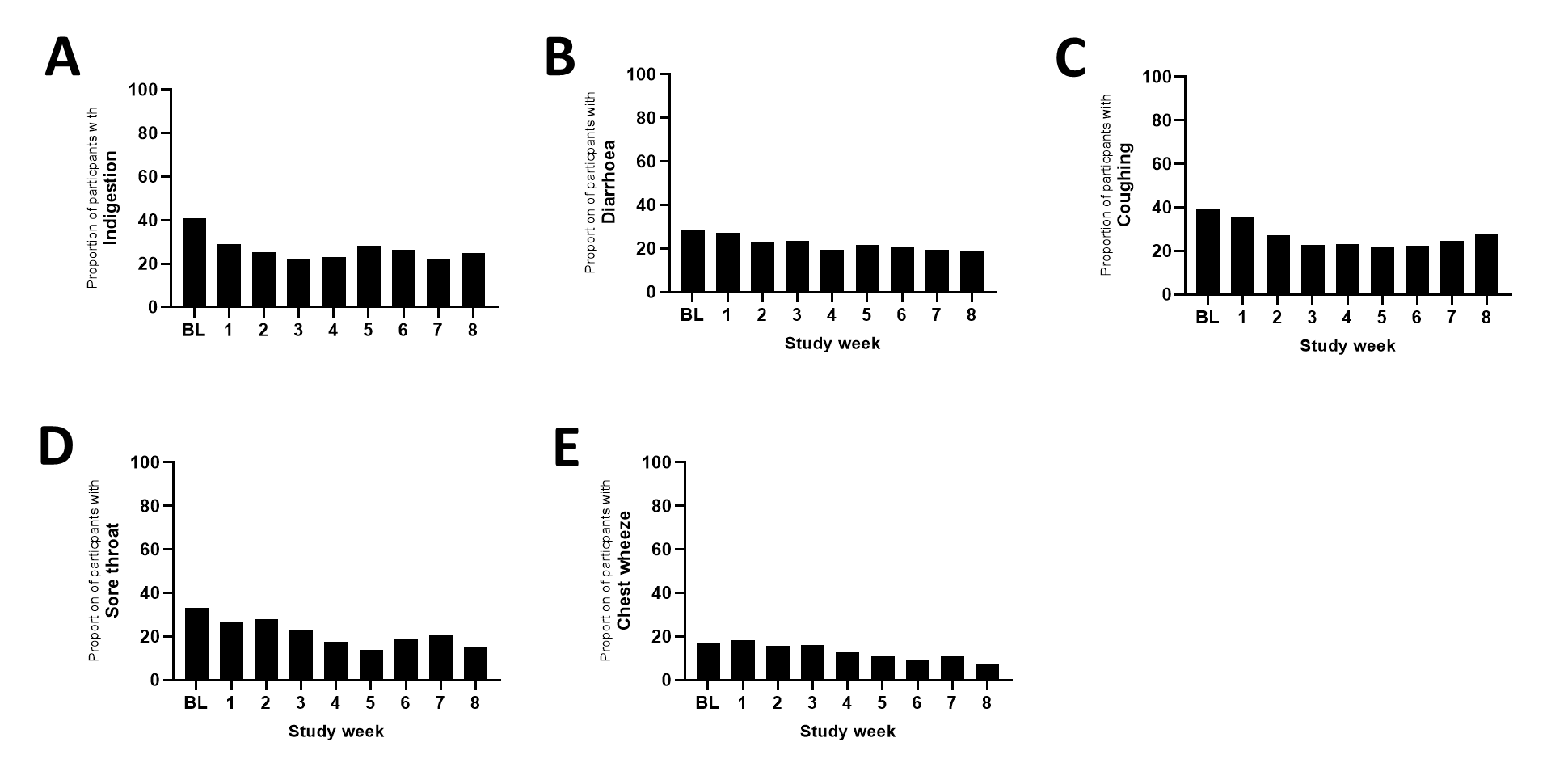

**Supplementary Figure S1.** **Incidence of participants reporting (A) Indigestion, (B) Diarrhoea, (C) Coughing, (D) Sore Throat and (E) Chest Wheeze over the duration of the study.** Data are presented as proportion (%) of participants reporting symptoms for n=159 at Week 1, n=147 at Week 2, n=137 at Week 3, n=125 at Week 4, n=110 at Week 5, n=102 at Week 6, n= 98 at Week 7 and n=97 at Week 8. Abbreviations: BL, baseline
